# Harnessing Electronic Health Records to Study Emerging Environmental Disasters: A Proof of Concept with Perfluoralkyl Substances (PFAS)

**DOI:** 10.1101/2020.12.10.20243501

**Authors:** Mary Regina Boland, Lena M. Davidson, Silvia P. Canelón, Jessica Meeker, Trevor Penning, John H Holmes, Jason H Moore

## Abstract

**Objective:** Environmental disasters are anthropogenic catastrophic events that affect health. Famous disasters include the Chernobyl and Fukushima-Daiichi nuclear meltdowns, which had disastrous health consequences. Traditional methods for studying environmental disasters are costly and time-intensive. We propose the use of Electronic Health Records (EHR) and informatics methods to study the health effects of emergent environmental disasters in a cost-effective manner.

**Materials and Methods:** An emergent environmental disaster is exposure to Perfluoralkyl Substances (PFAS) in the Philadelphia area. Penn Medicine (PennMed) comprises multiple hospitals and facilities within the Philadelphia Metropolitan area, including over three thousand PFAS-exposed women living in one of the highest PFAS exposure areas nationwide. We developed a high-throughput method that utilizes only EHR data to evaluate the disease risk in this heavily exposed population.

**Results:** We replicated all five disease/conditions implicated by PFAS exposure, including hypercholesterolemia, proteinuria, thyroid disease, kidney disease and colitis, either directly or via closely related diagnoses.

**Discussion:** Using EHRs coupled with informatics enables the health impacts of environmental disasters to be more easily studied in large cohorts versus traditional methods that rely on interviews and expensive serum-based testing. By reducing cost and increasing the diversity of individuals included in studies, we can overcome many of the hurdles faced by previous studies, including a lack of racial and ethnic diversity.

**Conclusion:** This proof-of-concept study confirms that EHRs can be used to study human health and disease impacts of environmental disasters and produces equivalent disease-exposure knowledge to prospective epidemiology studies while remaining cost-effective.

**KEY MESSAGES:**

- Electronic Health Records can be used for studying health effects of environmental exposures
- PFAS exposure - disease associations were mainly replicated using EHRs
- EHRs represent a cost-effective method to augment traditional epidemiology studies

## 1. BACKGROUND AND SIGNIFICANCE

### 1.1 Exposure to Environmental Disasters Impacts Human Health

Environmental disasters are catastrophic events that occur and are the result of human activity (i.e., anthropogenic) and often inadvertent. These differ from natural disasters, which are presumed to be non-human caused (e.g., hurricane, volcano eruption), and intentional acts such as nuclear bombings. Environmental disasters often affect human health directly. Famous environmental disasters include the Chernobyl nuclear meltdown in Ukraine on April 26, 1986. The resulting effects on human health included an increased rate of spontaneous abortions, including an excess loss of 400 male fetuses following first-trimester exposure, and sex-ratio disturbances observed throughout Northern Europe followed by increased rates of thyroid cancer ^1-5^.

Another devastating environmental disaster was the Seveso Herbicide plant explosion that occurred on July 10, 1972 in Seveso, Italy. This resulted in the highest known exposure to dioxin or 2,3,7,8-tetrachlorodibenzo-p-dioxin (TCDD) in residential populations ^6-9^. Spontaneous abortions (i.e., miscarriages) increased 67.7% following the Seveso plant explosion, in addition fathers exposed tended to have more female offspring then expected^6-9^. Chloracne (a dermatological condition) also resulted but not due to prenatal exposure. More recently, the Fukushima-Daiichi nuclear disaster occurred in Japan on March 16, 2011 resulting in increased exposure to radiation and other chemicals. Review of medical and health records revealed no observed change in the fetal loss or anomaly rate following the Fukushima-Daiichi nuclear disaster ^10^.

The typical epidemiology framework for studying environmental disasters includes conducting interviews of the exposed and affected populations, surveys to collect health outcomes, and in many cases review of health records. High-quality epidemiology studies use biomarkers of exposure, including serum concentrations of PFAS. However, these are expensive, typically costing over $100 per test and thereby limiting the sample size for study populations. In addition, there are many different species of PFAS (described further below).

### 1.2 PFAS Exposure: a Critical Environmental Disaster

A recent environmental disaster in the United States of America (US) involves perfluoroalkyl substances (PFAS). PFAS are anthropogenic persistent organic pollutants resistant to degradation and high heat and consist of different species of **F**luorinated **O**rganic **C**ompounds (**FOCs**). PFAS were used in a variety of products (and in some cases are still used although production in the US has ceased). Products include firefighting foams, non-stick cookware, dental floss, fire and chemical resistant tubing, and more (**Table 1**)^11^. Due to their chemical structure, PFAS have surfactant properties, and are resistant to degradation and high heat. The two most common PFAS species found in serum in the US are perfluorooctanesulfonate (PFOS) and perfluorooctanoate (PFOA) ^12,13^. The effect of PFAS on human health is only recently gaining awareness and therefore could be termed an emerging environmental disaster.

**Table 1.**
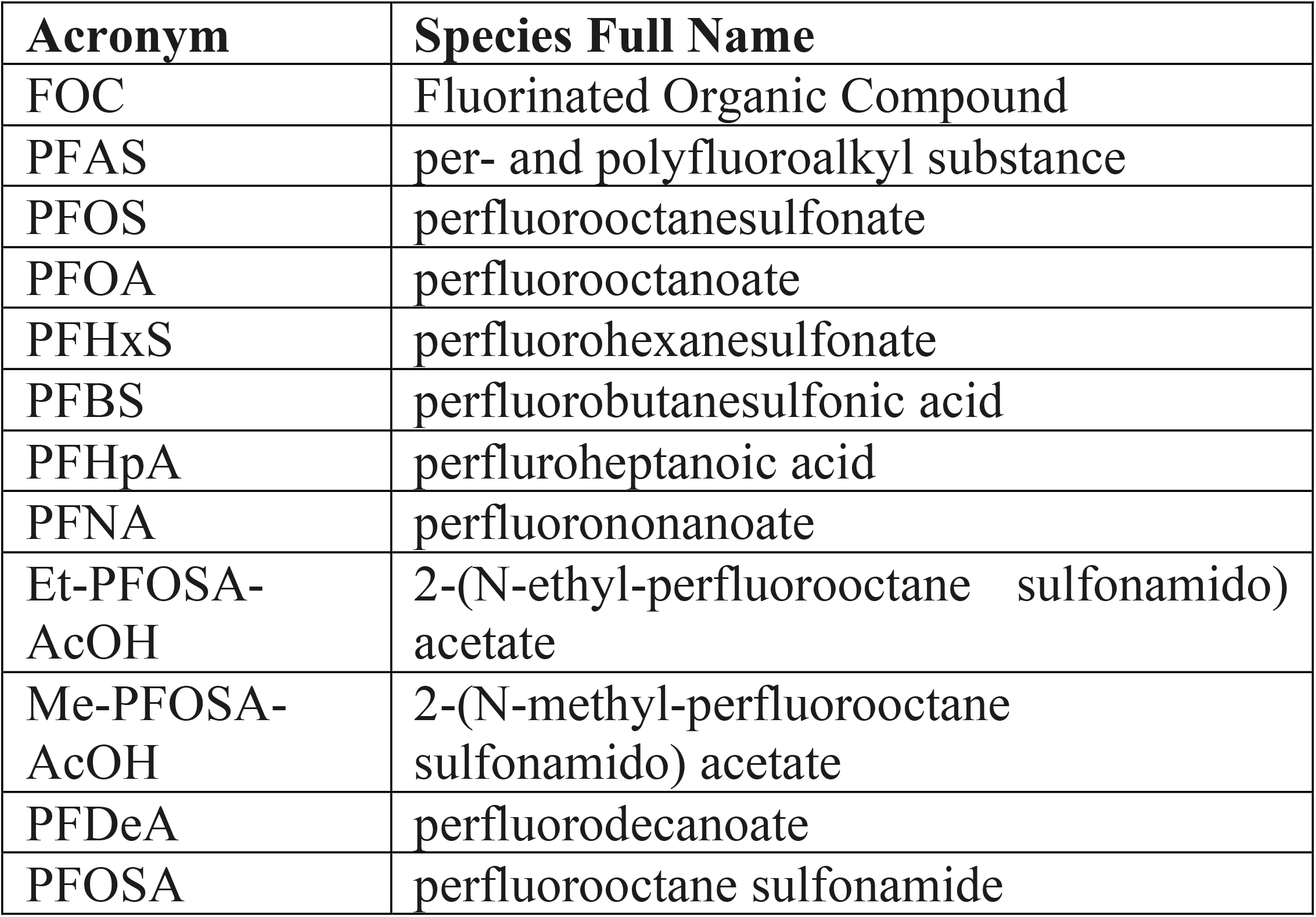
PFAS Species Names.

PFAS were detected in the Philadelphia area, specifically in Horsham-Warminster-Warrington following a series of events occurring in 2014 through May 19, 2016. In 2014, the established Provisional Health Advisory Levels (PHAL) were 200 ppt for PFOS and 400 ppt for PFOA (both types of PFAS). The Warrington Township Water and Sewer Department (WTWSD) promptly shut down public wells 1, 2 and 6 on October 29, 2014 (the same day they received the sampling results). Horsham Water and Sewer Authority (HWSA) took Well No. 26 and Well No. 40 off-line by August 2014, and three other wells had PFAS below the PHAL (HWSA Wells 10, 17 and 21). Warminster Municipal Authority (WMA) removed three wells

On May 19, 2016, US Environmental Protection Agency (EPA) released a new combined Health Advisory Level (HAL) for PFOS and PFOA of 70 ppt. WTWSD promptly shut down public wells 3 and 9 that very same day. HWSA shut down 3 wells in compliance. WMA removed another three wells, and started purchasing supplemental water supply from the Forest Park Water Treatment facility through the North Wales Water Authority (NWWA). Overall, three of the top 10 water utilities with the highest nationally recorded PFAS levels were located close to Philadelphia: Horsham-Warminster-Warrington ^14^.

PFAS exposure is known to increase the risk for various health outcomes, including high cholesterol^15^, ulcerative colitis^16,17^, thyroid disease^17,18^, kidney cancer^19^, pregnancy-induced hypertension, and preeclampsia^17,20^. In highly exposed US populations, indirect estimates of PFAS exposure were associated with preeclampsia ^21,22^, which puts these women at increased risk of cardiovascular disease (CVD) later in life^23^. Several observational studies have found PFAS exposure associated with high serum uric acid levels ^24-26^; epidemiologic evidence has linked elevated serum uric acid concentrations to hypertension ^27^.

### 1.3 Informatics Methods are needed to Harness Electronic Health Records for the Study of Health Impact

As mentioned previously, environmental epidemiologic studies typically rely heavily on interviews of the exposed and affected populations, surveys to collect health outcomes, recruitment of affected populations and in many cases manual review of health records. These studies require a painstaking amount of work and manual curation of specially designed datasets.

Data science has exploded these past few years with increasing numbers of studies seeking to incorporate ‘big data’ towards studying human health and disease. Electronic Health Records (EHRs) contain millions of patient records across billions of visits, including details such as laboratory values, medications, procedures, diagnostic imaging, clinical notes, and demographic data. Therefore, EHRs contain a treasure-trove of data that could be useful for studying the health impacts of environmental disasters, such as PFAS exposure in a given population, if the appropriate informatics methods are applied.

Previously, several disease-environment correlation studies have been performed using EHRs. The first was Patel *et al*. who coined the term EWAS: Environment-Wide Association Study ^28^. Others have studied environmental challenge in disease progression ^29^, thyroid cancer hotspots in a rural community in Vermont ^30^, and air pollution data in Italy ^31^. We have also conducted several studies investigating the relationship between birth season (as a proxy for seasonal variance in environmental exposures at birth) and disease risk ^32^. We conducted our initial study using New York City (NYC) data with novel findings that were validated in a separate EHR system ^33^. We expanded our study to include six sites including South Korea and Taiwan ^34^. This enabled us to develop a method that correlated birth season and trimester information with climate and pollution variables using location information ^34^. We have also used our methods to study the effects of social exposures that can manifest as birth month relationships for Attention Deficit Hyperactivity Disorder (ADHD) ^35^ and increased the number of datasets included in our assessment of female fertility and birth season to assess deeper and more complex multi-factorial environmental exposures ^36^. We have found that prenatal exposure to fine air particulates increased the risk of atrial fibrillation later in life in humans ^34^; this finding was supported by a study in a canine population ^37^.

These methods demonstrate that informatics methods can be developed to overcome EHR biases ^38,39^ and can be used to probe disease-environment interactions. However, they fall short in that they assess environmental exposures that are always present (e.g., air pollution) rather than the effect of an environmental disaster on human health and disease. An environmental disaster is more complex in that there is an exposure period, a remediation period and a post-exposure period. This differs from less-severe exposures that do not have a Government mandated remediation, clean up and post-remediation period.

This purpose of this study is to present a proof-of-concept detailing how EHRs can be used to study the impact of environmental disasters, such as PFAS exposure. We will compare the results generated from our EHR-only study to those that resulted from carefully curated epidemiology and government-based studies to demonstrate that we can effectively capture the health impacts of environmental disasters using EHR data only.

## 2. MATERIALS AND METHODS

This study involves the harmonization of data from the EPA, state and local water utilities and the Centers for Disease Control and Prevention (CDC) to appropriately capture the exposure – PFAS drinking water exposure. Once the exposure is properly categorized and assigned to a given latitude/longitude locations, we can correlate exposure with health outcomes using EHR data. We will describe these steps in detail below.

### 2.1 Integrating Diverse Data Sources to Define PFAS ‘Contaminated Regions’

The PFAS exposure in Horsham-Warminster-Warrington, PA was via the water supply. However, these three towns are located outside of Philadelphia city proper and therefore, some homes are on city/town water line while others are on private water wells. Exposure via the public water utilities (city/town water) is shown in **Figure 1**.

**Figure 1.**
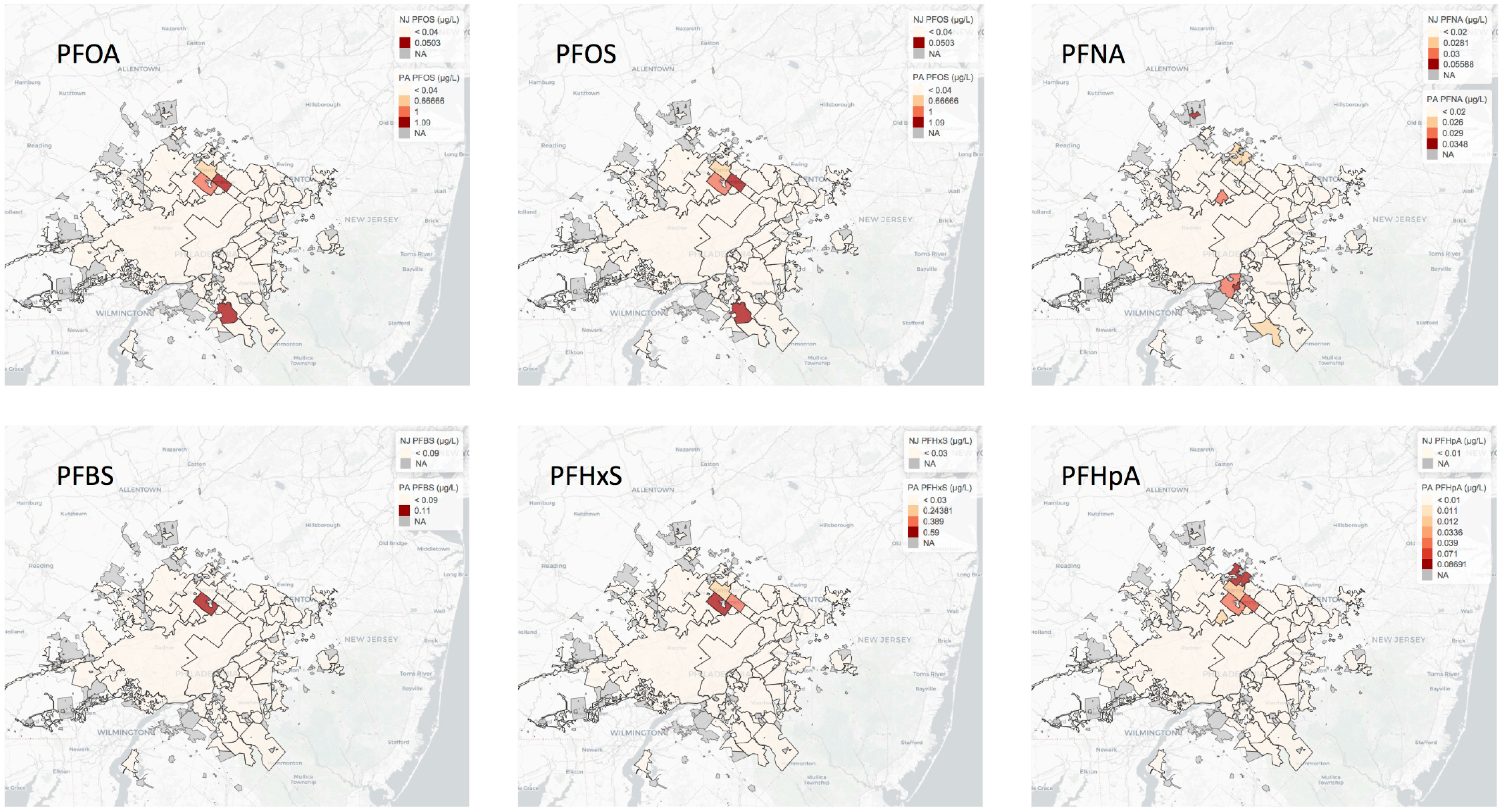
PFAS Exposure Levels as Reported in Public Water Utilities. Results over the Minimum Reporting Level (MRL) defined by UCMR-3 are recorded in µg/L and null values indicate results less than the MRL.

#### City/Town Water

Information and data on PFAS contamination of public drinking water supplies are available via the Third Unregulated Contaminant Monitoring Rule (UCMR-3) database maintained by the EPA ^40^. UCMR-3 monitoring occurred from 2013-2015 and included 6 PFAS species (PFOA, PFOS, PFNA, Perfluorobutanesulfonic acid (PFBS), Perfluoroheptanoic acid (PFHpA), and PFHxS) for assessment monitoring using EPA-approved analytical methods. While these 6 PFAS species do not represent all possible PFAS species (**Table 1**), they are the most common and EPA provides this information freely to the public. Public Water Systems have a unique identification code (PWSID) and may contain several facilities at which sampling occurred.

Boundaries of current public water suppliers’ (PWS) service areas are available from the PA Department of Environmental Protection (PA-DEP) Open Data Portal, which provides access to public non-sensitive GIS data^41^. The boundaries are approximate. The New Jersey Department of Environmental Protection, Water Supply and Geoscience^42^ maintains the water service area boundaries from the 2017 GIS data layer and includes systems that pipe drinking water to at least 15 service connections year-round, or systems that regularly serve at least 25 year-round residents. The boundaries are the actual water delivery or service area.

#### Well-Water

Over one million domestic water wells are distributed across Pennsylvania. The Pennsylvania Groundwater Information System (PaGWIS) contains information on well water and springs^43^. Data in PaGWIS are from completed water well reports collected since the mid-1960s and deposited in a database since the 1980s. In addition, 55,000 records contained in PaGWIS are field-located wells recorded in the US Geological Survey (USGS) database^43^. The PaGWIS contains information on well abandonment allowing us to identify locations of abandoned private wells. When a well is abandoned the reason is reported as either: a.) PFAS contamination, b.) poor water quality or c.) other reason. We are able to use this information along with the reported date of well-abandonment to determine if it is likely that the well was abandoned due to PFAS contamination (i.e., well was abandoned within six months of a reported PFAS water exposure in that area). Unfortunately, this dataset does not contain information on the PFAS species (**Table 1**) or other contaminants found in the well. Also not all well-testing information will be made available and time between testing may vary.

### 2.2 Linking Individual-Level Exposure to PFAS ‘Contaminated Regions’ with EHRs

For the purposes of this proof-of-concept study, we focused on women living in the three towns that were exposed to extremely high levels of PFAS. These towns are Horsham, Warminster and Warrington ^14^ referred to as Horsham-Warminster-Warrington throughout this study. These towns receive water supply from three of the top 10 water utilities with the highest nationally recorded PFAS levels in the entire US ^14^. Using address information contained in the EHRs, we linked patients to their precise geolocation within the Horsham-Warminster-Warrington area for visualization purposes. We used zip code information for the three towns, Horsham-Warminster-Warrington, to extract patients in exposed regions for our association analyses. We then extracted all diagnosis information contained in International Classification of Diseases (ICD) coding schema versions 9 and 10 for data collected between 2010-2017. These data were obtained during routine clinical care and stored in EHRs. The first PFAS exposure was detailed on May 19, 2016 and remediation occurred subsequently. We have data through 2017 and therefore all data is either pre-remediation or during the beginning of the remediation period. The biological half-life for PFAS species (shown in **Table 1**) ranges from 2 to 10 years depending on the compound^44^: 2-4 years for PFOA, 4-6 year for PFOS, and 8-10 years for PFHxS ^44^. Therefore, all patients with EHR data between 2010-2017 found in this heavily PFAS-exposed region would be expected to have elevated PFAS levels.

### 2.3 Statistical Analysis: Determining if Known PFAS Exposure-Disease Associations are Captured Using EHR clinical data alone

First we split our data into inpatient and outpatient records. We report the results obtained from the analysis of the inpatient records as these are less subjected to data missingness. We then performed Fisher’s exact test to determine the association between disease/condition/symptoms codes having at least 100 patients diagnosed at PennMed. WE compared the disease/condition/symptoms frequencies between those heavily exposed to PFAS (i.e., living in the Horsham-Warminster-Warrington towns) versus the overall cohort of patients treated at PennMed. Our approach was not ‘hypothesis-free’ in that we focused on diseases that were known to be related to PFAS exposure. This allowed us to confirm if EHRs are able to capture the known PFAS-disease relationships established in the epidemiology literature. These known PFAS-disease associations were established using carefully curated epidemiology studies and therefore they serve as a ‘gold-standard’. Associations have been established between high PFAS exposure and high cholesterol^15^, ulcerative colitis^16^, thyroid disease^18^ and kidney cancer^19^. Importantly, this research is mainly with PFOA and PFOS rather then all PFAS species found in the Horsham-Warminster-Warrington area. We will determine if we can effectively replicate all known PFAS-disease associations reported in the literature. We are excluding associations that are related to pregnancy (preeclampsia) as this requires capture of the pregnancy timeline. In addition, we are excluding testicular and prostate cancer, as our population is restricted to women.

The University of Pennsylvania’s Institutional Review Board approved this study.

## 3. RESULTS

### 3.1 EHRs can Capture Patients Exposed to Environmental Disasters such as PFAS

Using the zip code for individuals who were treated at Penn Medicine, we identified those living in Horsham-Warminster-Warrington. All of these women were included in our association analysis (results shown in **section 3**.**2)**. For mapping purposes, we linked address information recorded in the EHR to latitude and longitude. We identified the two major contamination sites in the Horsham-Warminster-Warrington area (depicted as gray shapes in **Figure 2**), along with the private well data denoting wells where PFAS contamination was reported for those exposed to the heavily polluted areas of Horsham-Warminster-Warrington (**Figure 2**). Contaminated wells are denoted with red circles in **Figure 2**.

**Figure 2.**
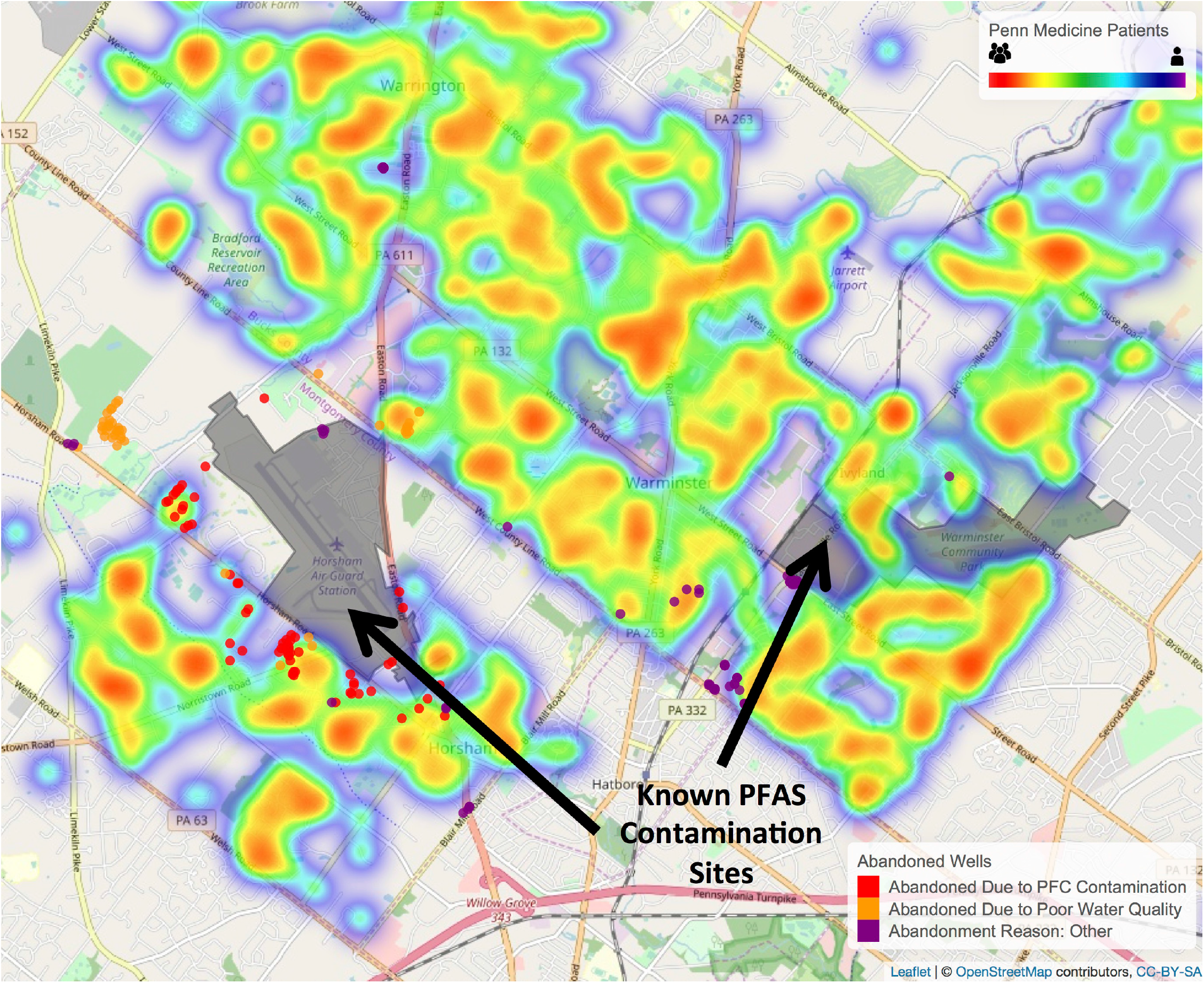
Heatmap detailing a subset of PennMedicine patients living in Horsham-Warminster-Warrington, PA along with PFAS contamination sites and water wells.

### 3.2 Validation of known disease-PFAS associations using clinical data from EHRs only

Women living the Horsham-Warminster-Warrington are exposed to PFAS at high levels. Therefore, we investigated whether or not we could replicate diseases known to be associated with high PFAS exposure among women visiting PennMed that lived in Horsham-Warminster-Warrington between 2010 and 2017 (during the highly exposed period). We investigated all diseases known to be elevated among women exposed to PFAS and compared the disease incidence to the background population. We found that high cholesterol, colitis and thyroid diseases (including multiple thyroid conditions) were elevated among women living in Horsham-Warminster-Warrington (**Table 2**). We also found the proteinuria was elevated, which is related to the elevated uric acid reported in the literature. We did not find kidney cancer to be elevated among these women, but found that complications related to kidney transplants were elevated, which could be indicative of prior kidney cancer (given that if kidney cancer was present previously then the diseased kidney may have been replaced with a transplanted kidney). We used Steenland *et al*.’s comprehensive review^45^ to determine ranges of odds ratios (ORs) reported in literature for diseases linked with PFAS exposure. If multiple studies were found, we took the study with the lowest reported OR, which for high cholesterol was 1.38 ^45,46^. There are reports of cardiovascular disease and cerebrovascular disease increases among PFAS exposed but these studies were too small for general conclusions according to Steenland ^45^. This review found no relation between kidney function and PFAS exposure; however, another study ^19^ reported a hazard ratio of 1.10 for kidney cancer for every 1 unit increase in PFOA in serum. Therefore, we looked for any kidney-related diagnoses that were nominally associated with PFAS exposure and found complications of transplanted kidney, which could indicate some impairment of kidney function among heavily exposed.

**Table 2.** Association Results for Known PFAS Exposed Patients (Horsham-Warminster-Warrington) and Diseases Known to be Associated with PFAS.

| Disease | Odds Ratio from Lit. | Ref | Disease in EHR | Odds Ratio from EHR (95% CI; P-Value) |
| --- | --- | --- | --- | --- |
| High Cholesterol | 1.38 | <sup>15,46</sup> | Pure hypercholesterolemia | 1.59 (CI=1.093, 2.237; p=0.013) |
| Elevated Uric Acid | 1.00-1.47 | <sup>45</sup> | Proteinuria, unspecified* | 6.67 (CI=1.351, 20.070; p=0.011) |
| Thyroid Disease | 1.64-1.86 | <sup>18,45</sup> | Malignant Neoplasm of the Thyroid Gland | 3.23 (CI=1.651, 5.706; p<0.001) |
|  |  |  | Primary Hyperparathyroidism | 3.93 (CI=1.061, 10.250; p=0.021) |
|  |  |  | Unspecified Acquired Hypothyroidism | 1.53 (CI=1.177, 1.967; p=0.002) |
| Kidney Cancer** | Hazard Ratio Reported | <sup>19</sup> | Complications of Transplanted Kidney** | 3.11 (CI=1.130, 6.851; p=0.015) |
| Ulcerative Colitis | Adj. Rate Ratio 1.76-2.86 | <sup>16</sup> | Toxic Gastroenteritis and colitis | 4.41 (CI=1.188, 11.502; p=0.014) |
|  |  |  | Regional Enteritis of Unspecified Site | 3.11 (CI= 1.411, 5.985; p=0.003) |

\*proteinuria and elevated uric acid are related but not the same (see Discussion)
\*\*Kidney Transplants Occur in Kidney Cancer Patients

Overall, these results demonstrate that clinical outcomes known to be due to high PFAS exposure in the literature are extractable from EHRs. In addition, it demonstrates that EHRs can be used to study the impact of environmental disasters on human health and disease without the need for specialized study recruitment and surveys.

## 4. DISCUSSION

Environmental disasters are large-scale anthropogenic events often with a fixed date - which for PFAS in the Horsham-Warminster-Warrington area reportedly ranges from October 29, 2014 to May 19, 2016. We demonstrate that EHRs can be used to study health outcomes following exposure to environmental disasters. Below, we discuss the many strengths of using EHRs to study environmental disasters and pollution in general over that of traditional epidemiology studies that require prospective patient recruitment.

### 4.1 Power of EHRs for Studying Impact of Environmental Disasters: Reduced Cost Enables More Exposed Populations to be studied

Epidemiology studies focused on environmental disasters – such as PFAS exposure in Horsham-Warminster-Warrington are expensive and often limited to a particular disease area, such as cancer. Government agencies, such as the EPA and the Agency for Toxic Substances and Disease Registry (ATSDR), often need to perform public health assessments on hazardous sites facing designation as a Superfund or Brownfield site. However, these public health agencies face many resource and financial constraints when deciding what outcomes to study, the wash out period following exposure, and in which locations ^47^. This leads to intense frustration in communities if their locations are not selected for further study, or if the outcomes they are most interested in are not explored – especially if they are already noticing alarming signs of deteriorating health among their loved ones. Even when a community is selected for study, it can be incredibly difficult for community members to wait for the results of the assessment, let alone the long process during which EPA and ATSDR decide on removal and remediation efforts. We should point out that the CDC did not select the Horsham-Warminster-Warrington site for future study likely due to budgetary constraints. In this current funding environment, studies using less expensive methods by harnessing EHR data become imperative.

The power of using EHRs to study environmental disasters such as PFAS, is that a wide range of health outcomes can be relatively quickly explored in an EWAS-style manner ^28^. The purpose of this current study is to serve as a proof-of-concept to demonstrate that we can replicate the known diseases associated with PFAS exposure in gold standard epidemiologic studies. This is a first step towards further detailed high-throughput analyses on diseases that individuals are at increased risk for following PFAS exposure. Furthermore, we demonstrate that EHRs are a powerful tool that can be used by public health investigators who are interested in studying disparate health outcomes.

### 4.2 Power of EHRs for Studying Impact of Environmental Disasters: Increased Inclusion and Diversity

Socioeconomic analyses in the study of PFAS exposure and its subsequent effects on human health and disease are also important ^48^. In this proof-of-concept, we ignore the socioeconomic factors because our desire was to replicate prior research on the health risks associated with PFAS exposure and the prior research often ignores socioeconomic co-contributors. A meta-analysis found that with a doubling of income the exposure to PFAS increased on average 10-14% ^49^. This is contrary to the typical environmental justice hypothesis that higher income individuals have lower exposure to pollutants in general ^49^. Many theories exist to explain this counter-intuitive result; some conclude that it could be due to routes of exposures (e.g., dental floss) or perhaps a result of the way many prior studies were conducted.

However, traditional epidemiology studies often require patient recruitment and consent and therefore are likely to result in inequities in part because recruiting representative populations is challenging ^50^. Recruiting minority populations is especially challenging due to historical inequities and distrust ^50^. Using EHRs to study environmental disasters, such as PFAS exposure, would address many of these issues. With a ready availability of clinical data from the EHR, a representative sample of the exposed population (in terms of race/ethnicity/sex/gender/various income levels) can be derived. This can mitigate some of the data collection and case ascertainment challenges experienced by traditional epidemiology studies.

### 4.3 Statistical Methods with EHR data can enable better Assessment of Biases in Environmental Studies

Analyzing EHR data can enable elucidation of issues with specific datasets, for example if a clinical dataset is derived from one type of clinic (e.g., ‘oncology’) the quality of the health assessment for other diseases that are non-oncology related will be affected ^51^. These dataset stratification issues could result in a biased assessment of certain exposure-disease related findings. Much work has been done to assess the bias of EHR methods and their portability from one clinic to the next ^34,52-55^. However, many environmental studies consist of meta-analyses of results across different sites without exploring site-specific biases that may affect the results. These types of biases can be addressed using informatics methods applied to EHRs that contain rich phenotypic data that is not often available from epidemiology studies. This is another strength of using EHRs and informatics methods to study environmental disasters. In addition, we open a new area of research for informatics through the exploration of health effects following environmental disasters.

### 4.4 Our Findings Versus the Literature: PFAS and Health Risks

#### Hypercholesterolemia

Our results for hypercholesterolemia are very similar to the literature with and OR of 1.38 being observed in the literature ^15,46^ and 1.59 (p=0013) being observed in our study. We did not have access to the exact laboratory values and therefore we should be underpowered to detect smaller non-diagnosis level changes in cholesterol levels. However, the PFAS exposure level is very high among our population and this likely explains our ability to detect these findings even using diagnosis code information alone. This demonstrates the strength of using EHRs for studying the health effects of emerging environmental disasters such as PFAS exposure in Horsham-Warminster-Warrington area.

#### Proteinuria

We found that the diagnosis code for proteinuria, unspecified was associated with PFAS exposure and elevated uric acid was reported in the literature as showing increases when PFAS exposure increased with an OR of 1.47 at the highest exposure quintile ^45^. We found that the diagnosis of proteinuria, unspecified was associated with PFAS exposure with an OR=6.66. Of note, we do not have the laboratory value results in this analysis and therefore, we are relying on the diagnosis code for ‘proteinuria, unspecified’ which is slightly different then elevated uric acid (**Table 2**). Importantly, it was noted that higher exposure levels showed higher uric acid and our location in Horsham-Warminster-Warrington contains three of the top 10 most contaminated water utilities in the entire US. Therefore, we likely have one of the most contaminated populations for PFAS exposure in the country, which may explain this very high risk of proteinuria observed.

#### Thyroid Disease

Previous studies have found links between various thyroid hormonal levels and PFAS exposure. However, because we do not have access to thyroid hormone laboratory results, we used the presence/absence of various thyroid conditions. In the literature ORs were observed for women for thyroid disease (OR=1.64) and taking medications related to thyroid disease (OR=1.86) with slightly lower ORs for males ^45^. We found that malignant neoplasm of the thyroid gland was elevated among women with OR=3.23 (p<0.001) and primary hyperparathyroidism was also elevated with an OR=3.93 (p=0.021). We also found that unspecified acquired hypothyroidism was elevated with an OR=1.53 (p=0.002), which was closer in risk size to those reported in previous studies ^45^. Initially, it may seem conflicting that both hypothyroidism and hyperthyroidism condition codes demonstrated increased risk in our PFAS exposure sample. However, many individuals who are being treated for thyroid disease have either hypo- or hyper-thyroidism at various stages of their therapy while the medication levels are being titrated ^56^. This could be the reason for our findings, and future work involves including laboratory values and medication histories for more detailed exploration in these findings. Importantly, the extremely high OR for malignant thyroid cancer in our population OR=3.23 indicates the important difference between studying PFAS exposure as an occupation as in other studies versus our population, which was heavily exposed chronically over a long period of time.

#### Kidney Disease/Cancer

A comprehensive review found no relation between kidney function and PFAS exposure ^45^. However, due to another study ^19^, we looked for any kidney-related diagnoses that were nominally associated with PFAS exposure and found complications of transplanted kidney (OR=3.11), which could indicate some impairment of kidney function among heavily exposed.

#### Colitis

A study found an adjusted rate ratio of 1.76-2.86 for ulcerative colitis among PFAS exposed individuals ^16^. In our heavily exposed population, we found that toxic gastroenteritis and colitis had an OR=4.41 (p=0.014) and ‘regional enteritis of unspecified site’ had an OR=3.11 (p=0.003) indicated that gastrointestinal issues and colitis are at increased risk among heavily PFAS exposed individuals.

### 4.5 Limitations and Future Work

There are several limitations of our work. We investigated findings reported in the literature to determine if we could replicate these well-known findings. We successfully validated known PFAS-disease exposures. However, it is important to note that other environmental exposures can also cause many of these diseases. For example, Endocrine Disrupting Chemicals can disrupt the thyroid system (e.g., PCBs and Dioxins). Our primary purpose was to determine if EHRs were useful in studying environmental disasters - given our ability to replicate the known PFAS-disease relationships, we conclude that they are a useful and cost-effective method of studying the health impacts of environmental disasters. However, additional studies would be required to rule out other exposures and concomitant comorbidities that may play a role.

From an informatics perspective, the alignment between diagnosis, symptom and condition codes found in the EHR and the diseases reported in these prior studies may not be exact. Future work, includes our plan to link disease codes from ICD-9 and ICD-10 to higher-level disease categories using ontologies such as the Disease Ontology or the Human Phenotype Ontology to assist in linking these codes to exact disease entities (although many of these methods have other limitations). However, it is still likely that those entities may differ from what was reported in the literature by other researchers. Therefore, we also plan to expand our analysis to include laboratory values; however, this is challenging due to the many different laboratory-testing services that exist with different reference standards and the porting of information in the EHR usually occurs in PDF format. Therefore this is the subject of future work. We also plan on using medication histories to determine treatment status (especially important for diseases/conditions like hypercholesterolemia and thyroid disease).

In addition there is the limitation of address accuracy, we used patient zip code information - if this was incorrect then we may be misclassifying patients as cases. Also, we do not have information on whether or not patients were using bottled water or not - therefore, it is likely we are underestimating the true effect of the PFAS exposure on human health. In addition, we classified everyone living between 2010-2017 in the Horsham-Warminster-Warrington as being exposed - this might not be the case if someone moved to the area in 2017 after the remediation period. This limitation also would lead us to underestimate the true effect of this exposure on human health.

## 5. CONCLUSION

In conclusion, we were able to successfully validate many of the known findings related to PFAS exposure reported in the literature using our heavily exposed PFAS population located in Horsham-Warminster-Warrington. This demonstrates the utility for using EHRs for studying emerging environmental disasters such as PFAS exposure in Pennsylvania. The advantages of using EHRs are described including the greater access to diverse patient populations that may not typically sign up for a clinical trial or detailed occupational study. In addition, we are able to study patients for longer periods of time then are often available to researchers who enroll patients, which is another advantage of EHRs for this type of environmental research.

## Data Availability

This paper uses Electronic Health Records data (EHR) obtained from the PennMedicine health system. This data is not freely available or shareable due to patient privacy concerns. Therefore the 'raw' patient data is not shareable. However, we are sharing our results in this paper and our methods are described.

## ACKNOWLEDGMENTS

We thank the Perelman School of Medicine at the University of Pennsylvania for generous funds to support this project. We also thank Nebojsa Mirkovic, Mark Miller, Sunil Thomas, and Selah Lynch for assistance within the Penn Data Store and Penn Data Analytics Center.

## CONTRIBUTORSHIP STATEMENT

Conceived Study Design: MRB

Developed methodology: MRB, LMD, SPC, JM

Provided environmental advice pertinent to study problem: TP, JHH

Provided informatics advice pertinent to study problem: JHM, JHH

Wrote Paper: MRB

Reviewed, Edited, and Approved Final Manuscript: MRB, LMD, SPC, JM, TP, JHH, JHM

